# One-pot CRISPR diagnostics for the rapid detection of *Candida* species and antimicrobial resistance biomarkers

**DOI:** 10.64898/2026.09.01.26361059

**Authors:** Amy Heathcote, Maya Bird, Hyeri Jeong, Xena Li, Robert Kozak, Nicole E. Weckman

## Abstract

Antimicrobial resistance (AMR) is a growing public health threat, where drug-resistant *Candida* infections are associated with increased hospital costs, lengths of stay, and high mortality rates. Current diagnostic methods are slow and labor-intensive, causing delays in diagnosis and patient treatment. In this research, we propose a rapid CRISPR-based diagnostic for identifying *Candida* pathogens and antifungal resistance biomarkers in clinical applications. One-pot CRISPR assay designs are optimized for three clinically relevant *Candida* species, achieving attomolar sensitivity with synthetic DNA and validating performance on cultured isolates. Strategies are developed to engineer CRISPR gRNA to control one-pot reaction kinetics, enhancing discrimination of single nucleotide polymorphism (SNP) resistance mutations associated with azole and echinocandin antifungals. This research expands our understanding of the design of one-pot CRISPR reactions for sensitive detection of DNA, while establishing a foundation for faster, more precise AMR diagnostics to improve surveillance and outbreak management of fungal infections.

## INTRODUCTION

In light of the growing global health threat of antimicrobial resistance (AMR), the spread of untreatable fungal diseases is of increasing concern. *Candida* species are the leading cause of invasive fungal diseases and rank fourth for the most common cause of hospital-acquired bloodstream infections with high mortality rates ranging from 38% to 57%^1–3^. Reflecting their clinical importance, the World Health Organization has placed *C. albicans, C. glabrata,* and *C. parapsilosis* on their pathogen priority list as they are the most recovered clinical species^4–6^. Azole and echinocandin antifungal resistance have been increasingly documented in *Candida* species, where some cases have shown multi-drug resistance, further complicating treatment options^3,7–9^.

AMR can arise from single nucleotide polymorphisms (SNPs) that lead to amino acid substitutions^10^. Azoles are the most common class of antifungals used to treat *Candida* infections^6^, while echinocandins are the first line of treatment for patients hospitalized with severe illness, or patients with prior exposure to azoles^6,11^. Given that antifungal susceptibility varies between species, routine antifungal susceptibility testing is critical to ensure effective therapy for patients^12^, and new rapid and accurate AMR diagnostics are urgently needed^13^. Early detection and identification of the pathogen and presence of antifungal resistance is essential for both effective treatment and limiting further spread in cases of transmissible pathogens.

Current diagnostic techniques to identify resistant species of *Candida* require specialized fungal cultures and micro-broth dilution-based assays; these are often performed only at reference laboratories with trained personnel, potentially delaying the start of effective therapy for the patient^7,14,15^. Critically, it has been shown that a 12-hour delay in initiating treatment has been linked to a minimum 20% increase in mortality rates from *Candida* bloodstream infections^13,14^.

Molecular diagnostics have the capability to detect DNA and RNA biomarkers within a few hours, offering a specific and sensitive approach for species-level detection^18^. Polymerase chain reaction (PCR) is a widely used method for highly sensitive and specific detection of nucleic acids^15,18,19^. However, PCR requires both cold-chain storage and thermocycler equipment, which limits its use in low-resource settings. PCR reagents are also susceptible to common inhibitors found in patient samples, and this method often lacks the specificity required to discriminate between SNPs, which is critical for AMR diagnostics^15,18–21^.

Clustered Regularly Interspaced Short Palindromic Repeats (CRISPR) based diagnostics are a class of molecular diagnostics that offers numerous advantages including cell-free, rapid, isothermal, and cost-effective results that can be tested directly at the point-of-care^15,22,23^. CRISPR-based diagnostics have been used to detect a number of pathogens including: SARS-CoV-2^24^, Zika Virus^21^, HPV^25^ and *M. tuberculosis*^26^. They have also been designed for SNP or single-variant detection^26–29^. However, challenges remain with CRISPR diagnostic design, including robustly detecting SNPs within various sequences, as well as translating CRISPR assays into streamlined one-step reactions that combine target amplification and CRISPR detection without loss of sensitivity or specificity.

To overcome the challenges of current AMR diagnostics, we propose a highly sensitive and specific CRISPR-based diagnostic for the detection of genetic biomarkers associated with *Candida* identification and antifungal resistance. We develop strategies to engineer gRNA designs with strategically placed mismatches with the target sequence that can control and improve reaction kinetics for streamlined one-step, isothermal assays. We show that these engineered gRNA can be used to manipulate Cas12a binding affinity to enhance SNP level specificity for AMR detection. Finally, we demonstrate the effectiveness of these techniques for achieving high-sensitivity and high-specificity detection of three common *Candida* species as well as a panel of associated antifungal resistance SNPs. These assays have the potential to enable rapid AMR diagnosis to support patient care and AMR surveillance efforts.

## RESULTS AND DISCUSSION

### *Candida* species detection

#### CRISPR-Cas12a two-pot assays

CRISPR-based diagnostics pair a sensitive isothermal amplification step like Recombinase Polymerase Amplification (RPA) with CRISPR-Cas12a recognition of a specific 20 bp dsDNA sequence for target or SNP identification (Figure 1A). Cas12a detects dsDNA target via designing a 20 bp guide RNA (gRNA) to bind a complimentary target sequence in the presence of an upstream PAM site (5’-TTTV-3’). Upon target recognition by the first 5 nucleotides in the PAM-proximal region, the Cas12a-gRNA complex is activated and cleaves both strands of dsDNA target, termed *cis*-cleavage^30,31^. This enables subsequent *trans*-cleavage of other nucleic acids, including spiked ssDNA reporter molecules, separating a fluorophore from a quencher to generate a fluorescent signal^15,30,32^.

**Figure 1.**
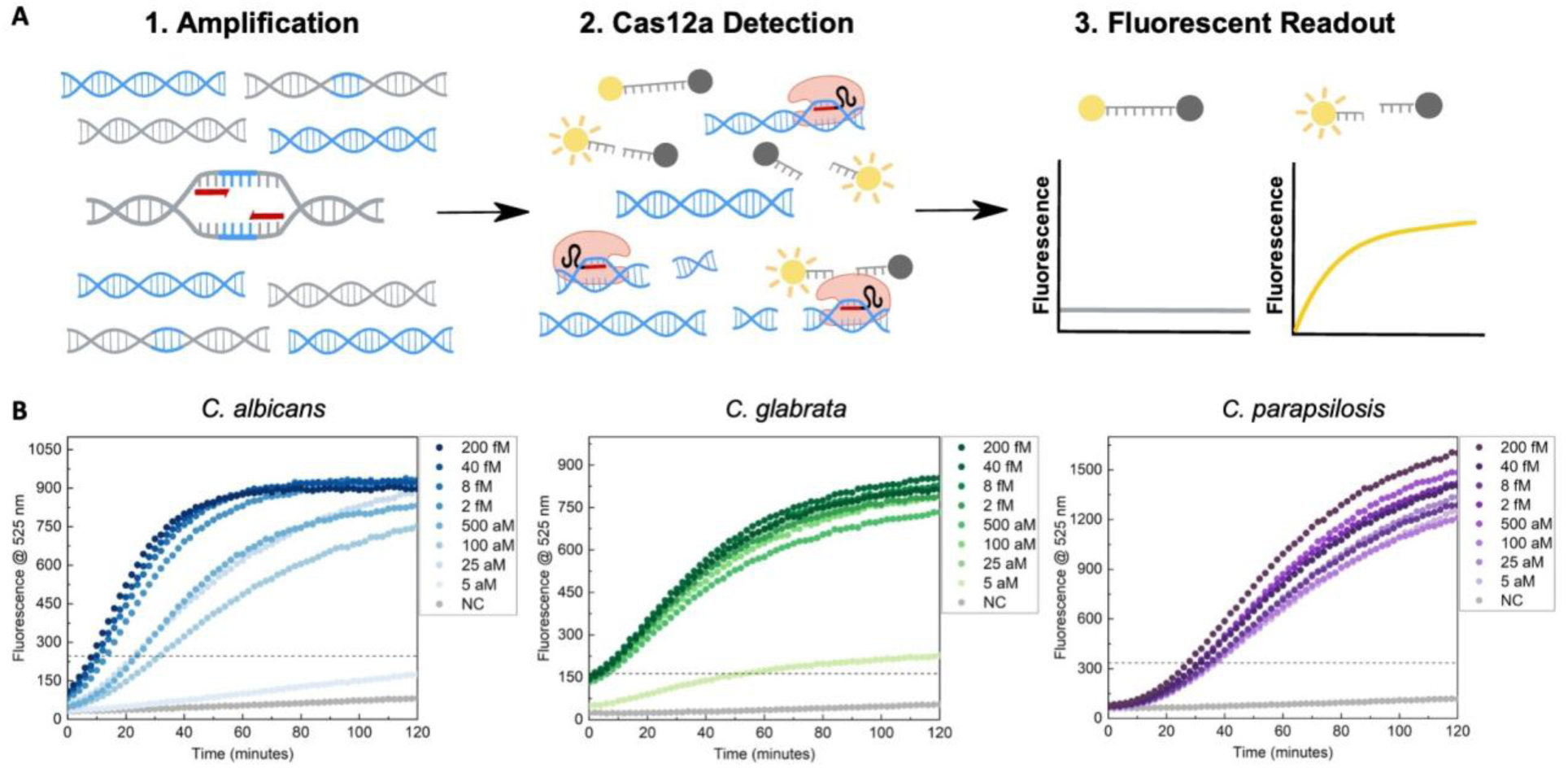
CRISPR-Cas12a two-pot sensitivity assay for *Candida* detection. (A) Schematic of a two-pot CRISPR-based diagnostic. Highly specific CRISPR-Cas nucleic acid detection is achieved by recognition of a target sequence by a programmable guide RNA (gRNA). The diagnostic workflow involves three key steps. (1) Target sequences (blue) are amplified by Recombinase Polymerase Amplification (RPA) to enhance sensitivity. (2) Cas12a-gRNA complexes (orange) bind to the amplified dsDNA and activate cis-cleavage of the target sequences. (3) Release of the 3’ end of the cleaved target sequence allows for trans cleavage activity of nearby quenched ssDNA reporter molecules, leading to a positive fluorescent signal. (B) Fluorescence intensity of *C. albicans* detection (blue), *C. glabrata* detection (green) and *C. parapsilosis* detection (purple) over 120 minutes. The negative controls (NC) without synthetic gBlock are in grey. Reference lines confirming detectable signal were defined by 3 times the negative control at 120 minutes.

In our initial efforts, we developed CRISPR-Cas12a assays to detect *C. albicans, C. glabrata,* and *C. parapsilosis.* These are the most common clinical species and are relevant for the surveillance and management of fungal infections, given their high incidence and increasing antifungal resistance profiles^6,33,34^. Highly conserved sequences within non-coding *ITS* regions were selected as target sequences for gRNA and RPA primer design^13,16,35–38^ (Table S1). Further details can be found in the Methods.

To select the RPA primer and gRNA pair that produced the highest and most rapid signal, screens were first performed in two-pot (*Step 1*: preamplification, then *Step 2*: Cas12a detection) (Figure 1B). The selected combinations for RPA preamplification and Cas12a detection were ITS2_3 gRNA with forward primer 8/reverse primer 7 (F8R7) for *C. albicans,* ITS2_1 gRNA and F4R4 for *C. glabrata,* and ITS1_1 gRNA and F3R3 for *C. parapsilosis* (Figure S1). All three assays demonstrated the ability to detect concentrations of synthetic DNA targets down to 25 aM after 40 minutes in the two-pot assays (Figure 1B). After an hour, *C. glabrata* and *C. parapsilosis* were able to recognize 5 aM of initial target DNA. This performance is consistent with previously reported CRISPR-based diagnostics and meets sensitivity benchmarks for molecular diagnostic tools^15,35^. Detection at such low DNA concentrations is essential for detecting early-stage infections and timely disease management efforts^15,21^.

#### CRISPR-Cas12a one-pot assays

Although two-pot assays offer high levels of sensitivity and specificity, the two-step amplification and detection process results in more liquid handling, increased risk of nucleic acid aerosol contamination and a longer time-to-result^21,24,39^. To streamline the workflow and minimize turnaround time, two-pot assays were consolidated into one-pot format. *C. glabrata* and *C. parapsilosis* showed similar results to their two-pot reactions, reaching 100 aM and 25 aM sensitivity, respectively, after 2 hours (Figure 2A). Compared to two-pot results, *C. albicans* one-pot assay showed reduced sensitivity and inconsistent fluorescence at different concentrations, no longer capable of attomolar detection (Figure 2A).

**Figure 2.**
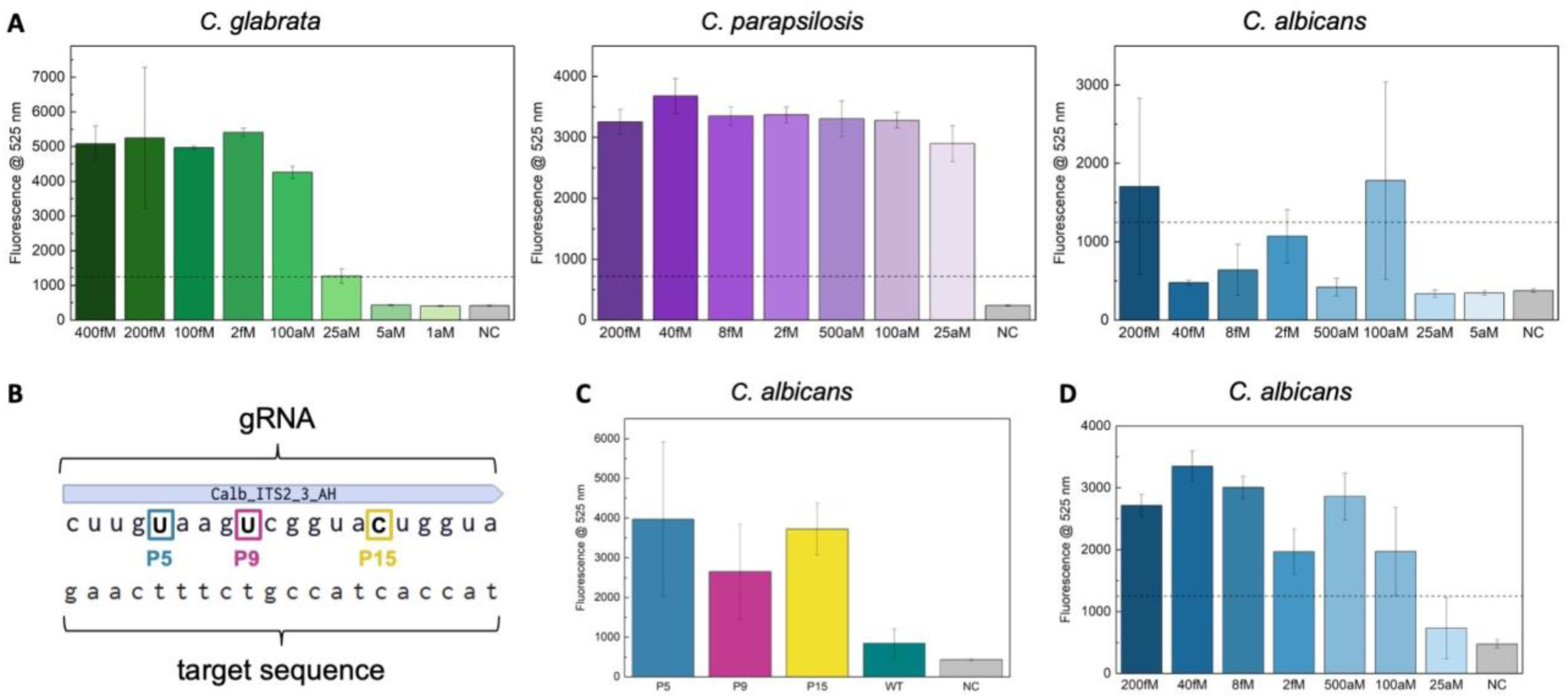
CRISPR-Cas12a one-pot assay optimization for *Candida* detection. (A) Fluorescence intensity of *C. glabrata* detection (green), *C. parapsilosis* detection (purple), and *C. albicans* detection (blue) after 120 minutes. The negative controls (NC) with no synthetic gBlock are in grey. (B) *C. albicans* engineered gRNA designs with synthetic mismatches at position 5 (blue), 9 (red) or 15 (yellow). (C) Fluorescence intensity of one-pot *C. albicans* gRNA screen comparing engineered gRNAs to wildtype 2_3 gRNA (green) after 120 minutes. (D) Fluorescence intensity of *C. albicans* detection with gRNA P5 and the same primer pair as in (A) and (C), after 120 minutes. The negative control (NC) with no synthetic gBlock is in grey. Reference lines confirming detectable signal were defined by 3 times the negative control at 120 minutes.

We hypothesized that the low sensitivity of the *C. albicans* assay is due to the Cas12a-gRNA complex outcompeting the RPA primers for target binding, resulting in cis cleavage and destruction of the target before amplification can occur^39–41^. Various strategies have been explored to mitigate this effect, including noncanonical PAM sites^28,42,43^, PAM-free gRNA^41,44,45^, engineered Cas12a^28,46,47^, and the addition of sucrose^39^, glycerol^48^ or heparin sodium^49^. In particular, engineering gRNA with synthetic mismatches is known to influence the binding kinetics and dissociation rates of Cas12a-gRNA complexes^30,31,50–54^ and has been shown to have the potential to improve one-pot sensitivity^29^. Based on these findings, we hypothesized that introducing synthetic mismatches along the gRNA could be used as a tool to control the relative cleavage and amplification kinetics, destabilizing the Cas12a-gRNA binding to the target sequence and delaying cis cleavage activity, thereby allowing more efficient RPA target amplification.

Synthetic mismatches were introduced at positions 5 bp (PAM-proximal), 9 bp, and 15 bp (PAM-distal) from the PAM to explore how the position of the mutation affects Cas12a cleavage activity and thus assay sensitivity (Figure 2B). PAM-proximal mismatches are known to have greater impact on gRNA binding kinetics than PAM-distal mismatches, where PAM-distal nucleotides are involved in complex stability^31,54–57^. For *C. albicans*, all engineered gRNA produced greater fluorescence in one-pot compared to the unmodified wildtype (WT) gRNA that perfectly matched the target sequence (Figure 2C). Engineered gRNA demonstrated faster reaction kinetics than the WT gRNA, with gRNA-P5 producing the earliest detectable signal at 48 minutes (Figure S2). These results highlight how mismatches at different positions can influence the binding and cleavage activity of the Cas12a-gRNA complex and align with previous studies demonstrating that Cas12a can be sensitive to all mismatches, regardless of their position^31,50^.

gRNA-P5, introducing a pyrimidine-pyrimidine synthetic mismatch at position 5, was selected for a one-pot sensitivity screen (Figure 2D). Using the engineered gRNA, attomolar-level detection was restored in the *C. albicans* assay, with detectable signal down to 100 aM after 2 hours. PAM-proximal nucleotides are key to target recognition and downstream enzyme activation^30,50^. In this case, the gRNA-P5 PAM-proximal mutation slows down complex binding to reduce rapid target destruction by Cas12a cis cleavage, enabling improved RPA amplification and thus restoring analytical sensitivity^30,50^.

#### Clinical Isolate Species Detection

To evaluate the specificity of the *Candida* CRISPR-Cas12a assays, we performed a specificity panel using heat-killed DNA to assess if any non-specific signal would occur across four other common *Candida* species (Figure 3). While PCR requires labor intensive sample preparation to remove potential RNA inhibitors, CRISPR-Cas12a assays are more robust, capable of sample detection after a simple lysis and 95 °C heat denaturation step for 30 minutes^15,23^. For all *Candida* species, these results were comparable to extracted DNA isolates (Figure S3), producing detectable signals within 1 hour. Overall, these results indicate that our diagnostic workflow can detect *Candida* species with high specificity in one-pot reactions and can be further streamlined by removing extensive sample purification steps, thereby reducing time-to-result.

**Figure 3.**
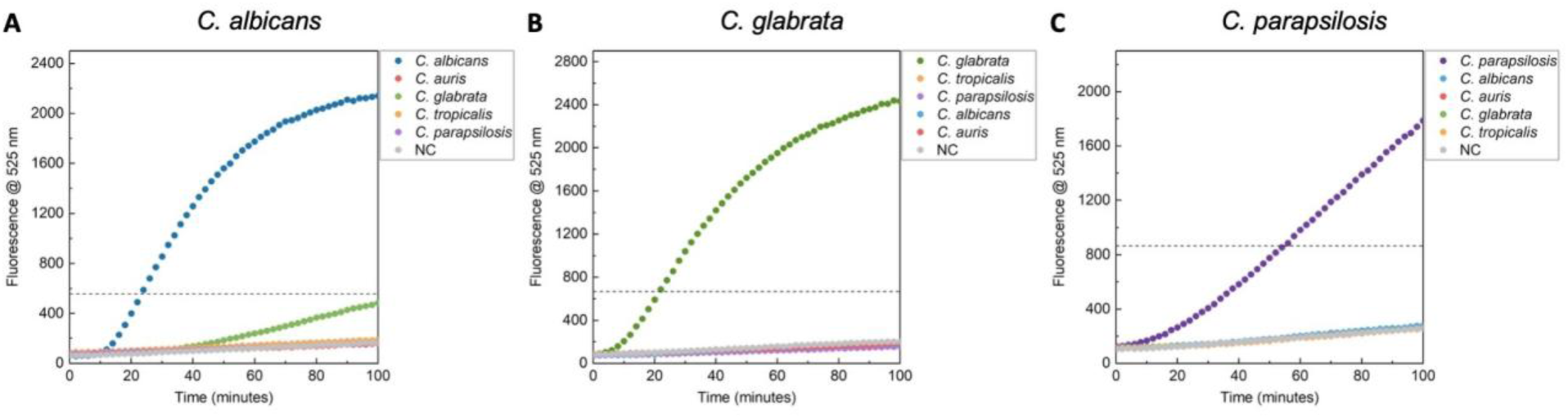
*Candida* clinical isolate CRISPR-Cas12a one-pot assays. (A) Fluorescence intensity of *C. albicans* detection (blue), (B) *C. glabrata* detection (green) and (C) *C. parapsilosis* detection (purple) over 100 minutes. *Candida* samples were from Shared Hospital Laboratory (Toronto, Canada) and heat-killed at 95 °C for 30 minutes. The negative control (NC) without genomic DNA is in grey. Reference lines confirming detectable signal were defined by 3 times the negative control at 120 minutes.

### Antifungal resistance detection

#### CRISPR-Cas12a gRNA Design for SNP Detection

In order to identify relevant SNPs for detecting AMR in *Candida* species, we performed a literature review to identify the most common mutations associated with resistance. We then prioritized designing assays for the most common AMR SNPs that are also adjacent to a PAM binding site to enable Cas12a binding (Table S2).

*C. albicans* and *C. glabrata* gRNA and primer sequences were designed to target the wildtype sequences corresponding to each resistance-associated mutation (Table S3). This strategy enabled the CRISPR-Cas12a complex to bind with perfect complementarity to the AMR hotspot regions in the absence of mutations, producing a strong fluorescence signal. If any nucleotide substitution was present, regardless of the specific amino acid change, it was expected to reduce the signal. This design helped mitigate against variability in the SNP mutation, where a positive fluorescence readout from our assay indicated no SNPs were present within a hotspot, suggesting the species is susceptible to the corresponding antifungal treatment (Table S4).

For *C. parapsilosis*, gRNA sequences were designed to target the sequence corresponding to a specific resistance-associated mutation (Table S3). In this case, the CRISPR-Cas12a complex binds complementary to the given mutation, allowing one to infer the presence of a specific mutated nucleotide. In contrast to the above, a positive fluorescence readout indicates that the target SNP is present, suggesting that treatment with the corresponding antifungal should be avoided (Table S4).

#### C. albicans ERG11 SNP detection

The *ERG11* F126 SNP is a known azole resistance hotspot in *C. albicans* and was targeted using gRNA 126, which was designed to bind complementary to the wildtype sequence (Figure 4A). We hypothesized that the SNP would prevent gRNA binding to the AMR sequence; however, results showed no difference in fluorescence signal between the wildtype and AMR mutant sequences, suggesting cross-reactivity of the unmodified gRNA (Figure 4B). This outcome aligns with previous findings stating Cas12a can tolerate multiple mismatches within its target sequence, limiting SNP discrimination^30,50,51^. To improve specificity we engineered gRNA based on the findings that Cas12a cleavage decreases significantly when two mismatches occur within 5 _bases32,50,58._

**Figure 4.**
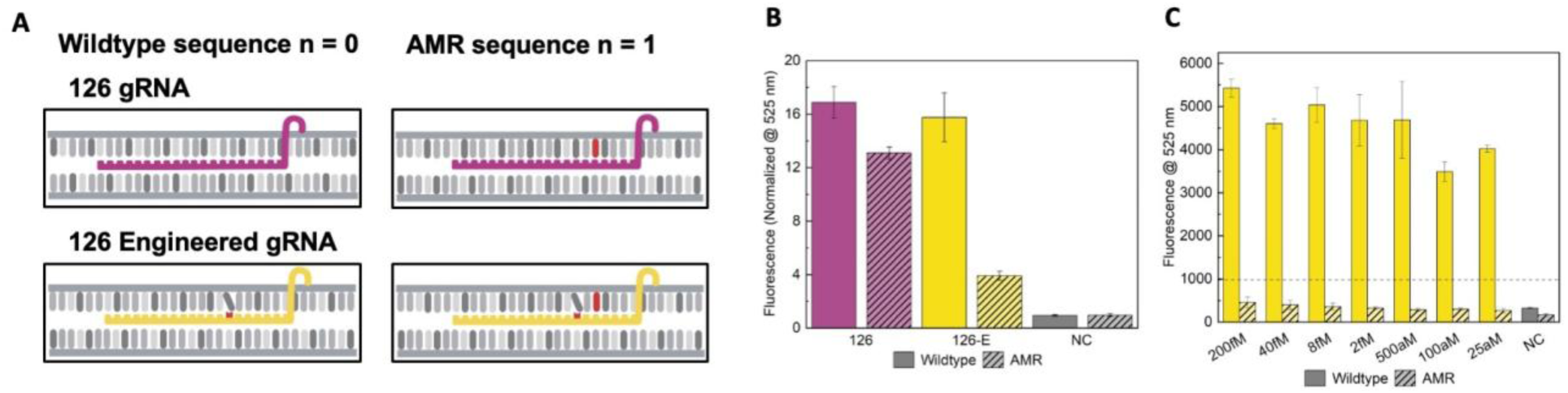
*C. albicans* 126 AMR hotspot detection in the *ERG11* gene. (A) 126 wildtype gRNA (pink) and 126 engineered gRNA (yellow) designs. The red nucleotide in the gRNA represents a synthetic mismatch with the shifted nucleotide, and the red nucleotide in the AMR sequence represents a SNP. (B) Fluorescence intensity of two-pot gRNA screen comparing 126 and 126-E gRNAs after 120 minutes. Fluorescence was normalized to t = 0. The negative controls (NC) without gRNA are in grey. (C) Fluorescence intensity of one-pot 126 AMR hotspot sensitivity screen with gRNA 126-E, after 120 minutes. The negative controls (NC) without synthetic gBlock are in grey. Reference lines confirming detectable signal were defined by 3 times the negative control at 120 minutes.

An engineered gRNA with a mismatch at position 7, two bases downstream of the SNP, was designed to improve differentiation of wildtype from mutant DNA (Figure 4A). The performance of engineered gRNA (126-E) and gRNA 126 were compared in a two-pot CRISPR-Cas12a assay (Figure 4B). Both gRNA produced comparable levels of fluorescence for the wildtype sequence, but 126-E showed significantly lower signal for the AMR sequence, demonstrating that the synthetic mismatch enhanced SNP discrimination. In a one-pot sensitivity screen, gRNA 126-E maintained high specificity and detected concentrations as low as 25 aM after 2 hours (Figure 4C). For *C. albicans,* we also designed an assay to detect an *ERG11* SNP at position 132. In this case, engineered gRNA 132-E was similarly able to discriminate between wildtype DNA and the AMR mutation, achieving attomolar sensitivity in a one-pot assay (Figure S4A-C).

#### C. glabrata FKS2 SNP Detection

The *FKS2* echinocandin resistance hotspot in *C. glabrata* contains only one PAM site near SNP positions 663 and 666. Within this region, three possible AMR variants have been identified: a SNP at position 663 only, a SNP at position 666 only, or the presence of both mutations simultaneously. This hotspot was selected for its clinical importance and because it provided a unique opportunity to determine whether a single gRNA could discriminate against all three AMR variants while still producing a detectable signal for the wildtype sequence^59,60^.

Three engineered gRNA were designed with synthetic mismatches (Figure 5A). When compared to the unmodified FKS2 gRNA, gRNA-E1 was the only gRNA to successfully discriminate against all three AMR variants (Figure 5B). Even if only 1 SNP was present, at either position, gRNA-E1 with 2 synthetic mismatches was needed to discriminate against the AMR sequences. These results suggest Cas12a has a higher SNP tolerance for *C. glabrata FKS2* target sequences (n = 3) than *C. albicans ERG11* target sequences (n = 2), suggesting Cas12a activity is influenced by the specific composition of the target sequence.

**Figure 5.**
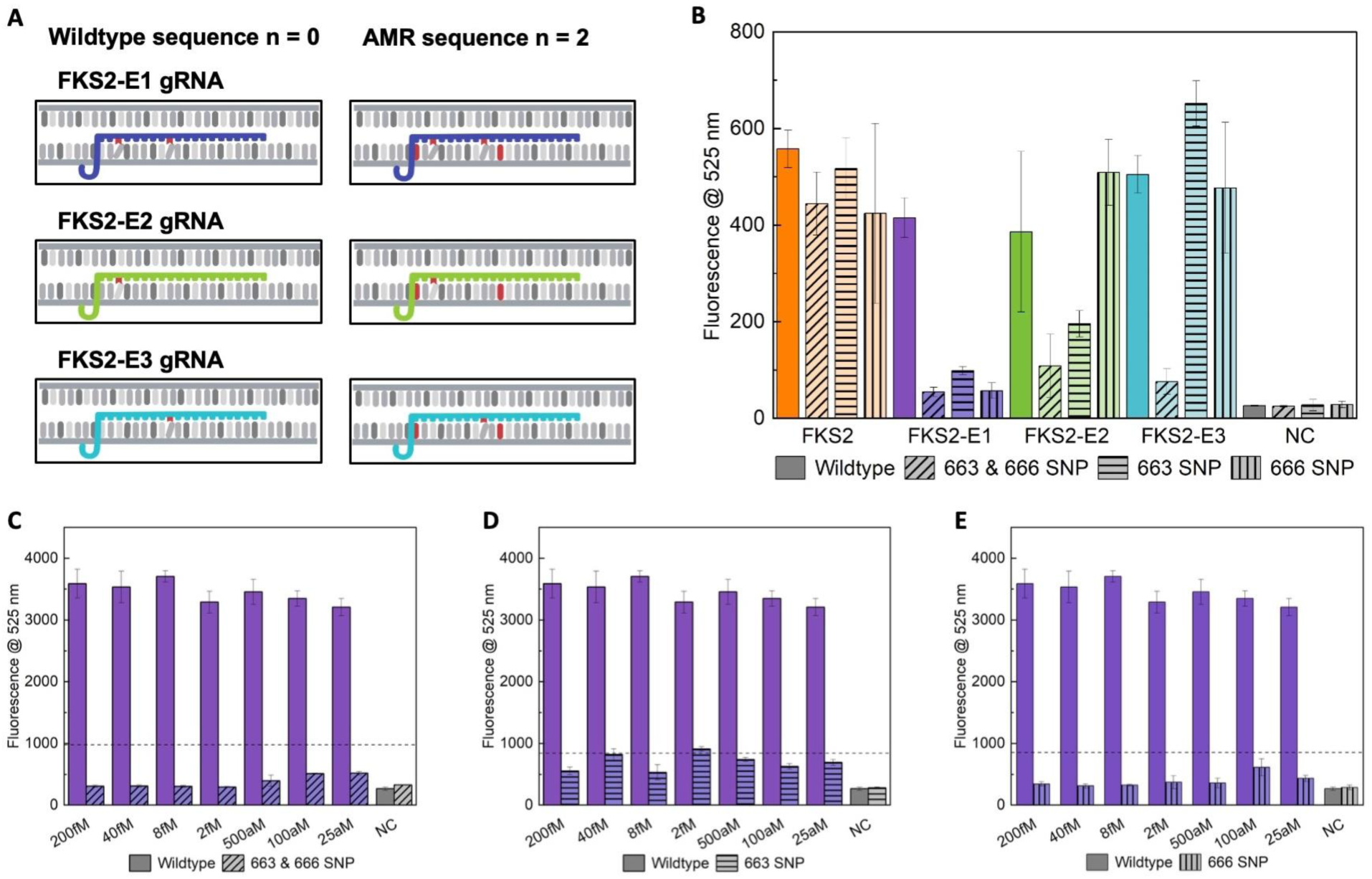
*C. glabrata* 663 and 666 AMR hotspot detection in the *FKS2* gene. (A) *FKS2* engineered gRNA designs. The red nucleotide in the gRNA represents a synthetic mismatch with the shifted nucleotide, and the red nucleotide in the AMR sequence represents a SNP. AMR sequence with 663 & 666 SNPs was used as an example. (B) Fluorescence intensity of two-pot gRNA screen comparing engineered gRNAs after 120 minutes. The negative controls (NC) without gRNA are in grey. (C-E) Fluorescence intensity of one-pot sensitivity screens with gRNA FKS2-E1, after 120 minutes. Wildtype target sequence detection compared to (C) AMR 663 & 666 SNP sequence, (D) AMR 663 SNP sequence and (E) AMR 666 SNP sequence. Negative controls (NC) without synthetic gBlock are in grey. Reference lines confirming detectable signal were defined by 3 times the negative control at 120 minutes.

When analyzing the fluorescence signals of FKS2 engineered gRNAs targeting the wildtype sequence, gRNA-E1 and gRNA-E2 produced lower signal than gRNA-E3, suggesting reduced Cas12a activity (Figure 5A). Mismatches in the PAM-proximal region have been reported to have the greatest effect on R-loop formation and cis cleavage activity^25,58^. Consistent with this model, gRNA-E1 and gRNA-E2 with synthetic mismatches at position 3 greatly reduced activity and further suppressed nonspecific signal from AMR variants when SNP 663 was also present at position 1 of the gRNA. In a one-pot CRISPR-Cas12a sensitivity analysis, gRNA-E1 was able to selectively detect the wildtype sequence against 3 different AMR variants, while achieving attomolar sensitivity (Figure 5C-E). These results highlight the potential of CRISPR-based diagnostics to rapidly and specifically discriminate against wildtype sequences from AMR variants with 1-2 SNPs in less than 2 hours.

#### C. parapsilosis FKS1 SNP detection

For detection of *C. parapsilosis FKS1* echinocandin resistance at position 656, an engineered gRNA was designed with a synthetic mismatch at position 11, two bases upstream of the SNP (Figure 6A). The specificity of engineered gRNA (656-E) and gRNA 656 were compared in a one-pot CRISPR-Cas12a assay (Figure 6B). Compared to *C. glabrata* target sequences, Cas12a was the least tolerant to PAM-distal mismatches adjacent to SNP 656 in *C. parapsilosis,* where a synthetic mismatch at position 11 in the gRNA substantially lowered fluorescent signal for both target and non-target sequences. PAM-distal mismatches have been shown to destabilize the Cas12a-gRNA complex, increasing dissociation rates and reducing target binding^31^. In this instance, the mismatch at position 11 and position 13 decreased Cas12a-gRNA binding kinetics, allowing for SNP discrimination and target sequence specificity to occur. Following a sensitivity screen, gRNA 656-E detected concentrations down to 25 aM, maintaining high sensitivity and specificity for the AMR sequence (Figure 6C). Engineered gRNA targeting AMR SNP 660 followed similar specificity trends for the AMR target sequence and achieved attomolar sensitivity in a sensitivity screen (Figure S4D-F).

**Figure 6.**
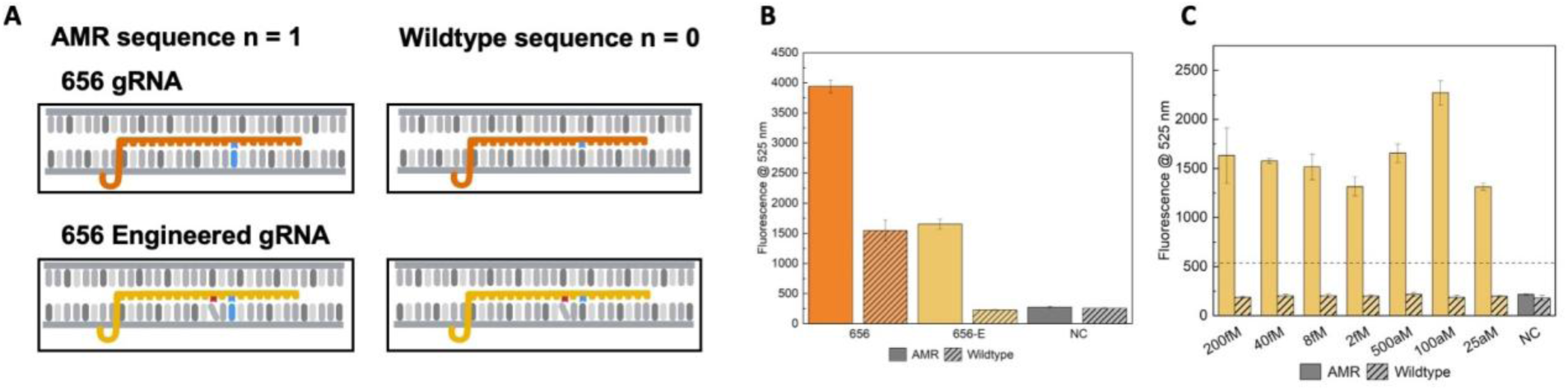
*C. parapsilosis* 656 AMR hotspot detection in the *FKS1* gene. (A) 656 wildtype gRNA (orange) and 656 engineered gRNA (yellow) designs. The red nucleotide in the gRNA represents a synthetic mismatch with the shifted nucleotide. The blue nucleotide in the gRNA represents complementarity with the SNP, represented by the blue nucleotide in the AMR sequence. (B) Fluorescence intensity of one-pot gRNA screen comparing 656 and 656-E gRNAs after 120 minutes. The negative controls (NC) without gRNA are in grey. (C) Fluorescence intensity of one-pot 656 AMR hotspot sensitivity screen with gRNA 656-E, after 120 minutes. The negative controls (NC) without synthetic gBlock are in grey. Reference lines confirming detectable signal were defined by 3 times the negative control at 120 minutes.

## CONCLUSION

The increasing threat of multidrug-resistant fungal pathogens underscores the need for rapid diagnostics capable of single-nucleotide specificity. We developed CRISPR-based assays that detect three clinically relevant *Candida* species with sensitivities below 100 aM in under 1 hour. In parallel, we established assays to detect SNPs associated with azole and echinocandin resistance mutations, achieving detection sensitivities of 25 aM in under 2 hours.

Our work highlights the challenges of designing one-pot CRISPR assays for SNP detection and clarifies the mechanisms governing gRNA specificity and Cas12a cleavage. Cas12a efficiency is highly sequence-dependent, with mismatches at multiple gRNA positions influencing enzymatic activity. Importantly, these findings demonstrate that gRNA engineering is critical to enhance analytical sensitivity and enable single-nucleotide specificity in one-pot assays, revealing the complex interactions between binding kinetics, amplification efficiency, and cleavage activity. Further studies are needed to systematically characterize Cas12a–gRNA–target interactions. Collectively, our results provide a framework for CRISPR-diagnostic optimization, supporting the use of gRNA screening with synthetic mismatches to restore sensitivity or specificity when performance is compromised.

Our final one-pot approach achieves a total turnaround time of approximately 3 hours, including sample preparation, while reducing processing steps and hands-on effort relative to traditional culture-based and PCR diagnostics. This streamlined approach positions our CRISPR assays as a valuable future tool for hospital clinical microbiology laboratories and reference laboratories, particularly during *Candida* outbreaks. By enabling rapid identification of antifungal resistance mutations, this approach has the potential to inform early therapeutic decisions and strengthen antimicrobial stewardship. More broadly, this research provides a framework for developing sequence-specific CRISPR diagnostics for clinically relevant pathogens.

## MATERIALS AND METHODS

### Target sequence selection

Highly conserved sequences within non-coding *ITS* regions unique to *C. albicans, C. glabrata* and *C. parapsilosis* were identified by previous studies^13,16,35–38^. For AMR detection, *ERG11 FKS1, and FKS2* genes associated with azole and echinocandin resistance were identified for each species, along with the most common SNPs that confer resistance (Table S2).

Sequences were retrieved from the NCBI GenBank ([NCBI GenBank](https://www.ncbi.nlm.nih.gov/genbank/)) and analyzed by sequence alignment against different strains and isolates to identify conserved regions and AMR hotspots suitable for gRNA and primer design. Species specific and AMR sequences were ordered as gBlock gene fragments from IDT ([IDT DNA](https://sg.idtdna.com/pages/products/custom-dna-rna/dna-oligos/custom-dna-oligos)).

### gRNA design and synthesis

gRNA were designed within target regions as described in Results. gRNA templates were ordered as ultramer ssDNA oligos ([IDT DNA](https://www.idtdna.com/pages/products/custom-dna-rna/dna-oligos/custom-dna-oligos)) with a T7 promoter region, structural gRNA sequence and 20 bp gRNA sequence. In vitro transcription of gRNA ultramers were performed using HiScribe T7 High Yield RNA Synthesis Kit (E2050S; New England Biolabs). gRNA were purified using the Zymo Research RNA cleanup kit (R1017, Zymo Research). All products gRNA were diluted to 100 µM and stored in −20 °C until use.

### CRISPR Sensitivity Assay

Fluorescence based assays were first designed in a two-pot reaction involving an RPA preamplification step and a subsequent Cas12a detection step. The preamplification step was completed as following the manufacturer’s instructions using the TwistAmpBasic Kit (TABAS03KIT, TwistDx). The Cas12a detection step was prepared using a master mix of 9.4X NEB Buffer r2.1 (Cat. No.: B6002S; New England Biolabs, Ipswich, MA, USA), 1.25 µM gRNA, 1.25 µM LbCas12a (Cat. No.: M0653T; New England Biolabs, Ipswich, MA, USA) and 4 µM FQ reporter molecule (56-FAM/TTATT/3IABkFQ). A 2.5 µL volume of master mix was added to the RPA product to make a final reaction volume of 52.5 µL. Samples were incubated for 30 minutes at 37 °C. Samples were mixed and transferred to a 384-well plate (Cat. No.: 3820; Corning Inc.). Fluorescence measurements were taken using a microplate reader (λ_ex_ = 493, λ_em_ = 525 nm, gain = 100).

### CRISPR One-Pot Assay

One-pot reactions were prepared uing two master mixes to precisely control the reaction initiation time. The RPA master mix was composed of 10 µL PEG (Tris [0.02], NaCl [0.06], PEG – 35k 0.05 w/v), 0.75 µM of each RPA primer and nuclease free water for a total of 33.3 µL in 1 TwistAmp Basic RPA pellet (TwistDx). The Cas12a master mix composed of 5.4X NEB r2.1 buffer, 2.7 µM gRNA, 0.27 µM LbCas12a, 6.8 µM FQ and also contained the 93 mM of MgOAc (TwistDx) needed to initiate the RPA reaction. A volume of 2 µL of gBlock, 6.5 µL of RPA master mix and 1.5 µL Cas12a master mix was mixed together and incubated at 37 °C for 15 minutes. Samples were transferred to a 384-well plate (Cat. No.: 3820; Corning Inc.). Fluorescence measurements were taken using a microplate reader (λ_ex_ = 493, λ_em_ = 525 nm, gain = 100).

### Clinical Isolate Preparation for Species Detection

*Candida* strains were from clinical patient samples collected during routine patient care and preserved in glycerol stocks at −80 °C at Shared Hospital Laboratory (Toronto, Canada). For experimental use, isolates were cultured and recovered on CHROMagar Candida Plus plates (Micronostyx, Canada). Following this, DNA was either extracted using the easyMAG platform (bioMerieux, France) or samples were heat-killed at 95 °C for 30 minutes.

### Data Analysis

For the purposes of these diagnostics, the aim is to distinguish between the absence of a target (negative control) and its presence, and so the results have been evaluated based on qualitative validation methods^21^. A threshold was set at three times the negative control signal after 120 minutes and signals greater than this threshold were confirmed as positive detection. Error bars represent the standard deviation (±1 SD) of each plotted mean.

## Data Availability

All data produced in the present study are available upon reasonable request to the authors

## ASSOCIATED CONTENT

There is an attached **Supporting Information** file that contains further experimental details.

## AUTHOR INFORMATION

### Author Contributions

AH and MB designed and performed experiments, analyzed the data, and wrote the manuscript. HJ performed experiments. XL and RK provided clinical samples, clinical expertise and advised on experimental design. NEW and RK conceptualized research and secured funding, advised on experimental design, methods, and data analysis, and contributed to manuscript preparation. All authors have had input and given approval to the final version of the manuscript.

## ACKNOWLEDGMENT

Clinical samples were processed and tested at Shared Hospital Laboratory, Toronto, Canada.

