## Supporting Information for "One-pot CRISPR diagnostics for the rapid detection of *Candida* species and antimicrobial resistance biomarkers"

**Table S1. gRNA and RPA primer designs for identifying *ITS1* and *ITS2* regions within *Candida* species.** PAM sites (highlighted in bold) correspond to the first four nucleotides of each sequence. gRNAs were designed in the RNA form of these sequences, made up of the 20 nucleotides downstream the PAM site. Primer sequences were designed as DNA with lengths between 30 – 35 bp long, and amplicons ≥ 100 bp. All sequences are listed in the 5’ to 3’ orientation.

| | |
|---|---|
| ***Candida albicans* sequences** | |
| ITS2 gRNA 2 | **tttg**gtgttgagcaatacgacttg |
| ITS2 gRNA 3 | **tttg**cttgaaagacggtagtggta |
| ITS2 gRNA 3 M5/P5 | **tttg**cttgTaagacggtagtggta |
| ITS2 gRNA 3 M9/P9 | **tttg**cttgaaagTcggtagtggta |
| ITS2 gRNA 3 M15/P15 | **tttg**cttgaaagacggtaCtggta |
| ITS2 gRNA 4 | **tttg**acaatggcttaggtctaacc |
| Forward primer 7 | ccggagggcatgcctgtttgagcgtcgtttc |
| Forward primer 8 | ttgcgccctctggtattccggagggcatgc |
| Forward primer 9 | ctttgaacgcacattgcgccctctggtatt |
| Reverse primer 7 | gacgttaccgccgcaagcaatgtttttggt |
| Reverse primer 8 | aagatatacgtggtggacgttaccgccgca |
| Reverse primer 9 | aggtcaaagtttgaagatatacgtggtgga |
| ***Candida glabrata* sequences** | |
| ITS1 gRNA 1 | **ttta**tcacacgactcgacactttc |
| ITS2 gRNA 1 | **tttg**agttaacttgaaattgtagg |
| Forward primer 2 | acgttgtgtttggtagtgagtgatactctc |
| Forward primer 4 | tctggtattcatgcctgtttgagcgtcatt |
| Forward primer 5 | ctttgaacgcacattgcgccctctggtatt |

| | |
|---|---|
| Reverse primer 3 | ttaatagagaagcttgcgctcgtgtcccac |
| Reverse primer 4 | gcgcaaacgagcagcagattaatagagaag |
| Reverse primer 5 | taatatcgcgcaaacgagcagcagattaat |
| ***Candida parapsilosis*** **sequences** | |
| ITS1 gRNA 1 | **ttta**atgtcaaccgattatttaat |
| ITS2 gRNA 1 | **tttg**gtgttgagcgatacgctggg |
| ITS2 gRNA 2 | **tttc**cactcattggtacaaactcc |
| Forward primer 1 | ctgcattttttcttacacatgtgtttttct |
| Forward primer 2 | gaaaactttgctttggtaggccttctatatggggc |
| Forward primer 3 | taggccttctatatggggcctgccagagat |
| Reverse primer 1 | ggaataccaaagggcgcaatgtgcgttcaa |
| Reverse primer 2 | gtgcgttcaaagattcgatgattcacgaatatct |
| Reverse primer 3 | cacgaatatctgcaattcatattacttatcgcatt |

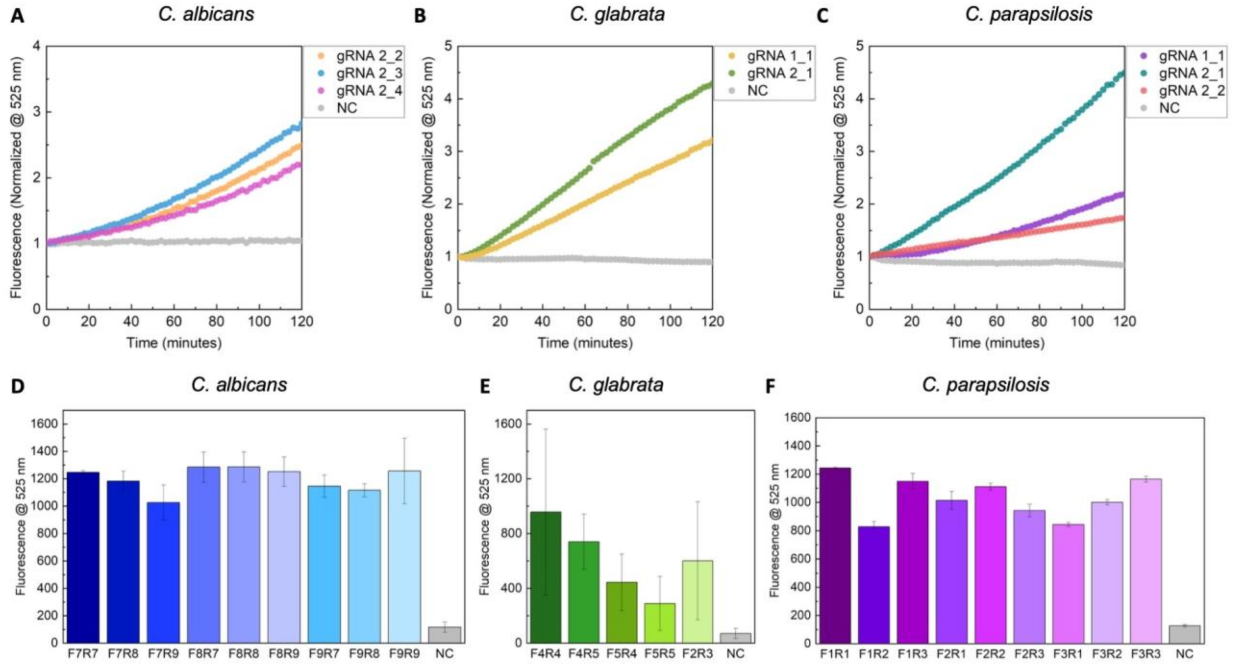


**Figure S1.** *Candida* CRISPR-Cas12a gRNA and RPA primer screens. (A) Fluorescence intensity of Cas12a gRNA screens for *C. albicans,* (B) *C. glabrata*, and (C) *C. parapsilosis* over 120 minutes. Fluorescence was normalized to t = 0. The negative controls (NC) without gRNA are in grey. (D) Fluorescence intensity of primer pair screens for *C. albicans* ITS2_3 gRNA (E) *C. glabrata* ITS2_1 gRNA, (F) and *C. parapsilosis* ITS1_1 gRNA after 120 minutes. The negative controls (NC) without primers are in grey.

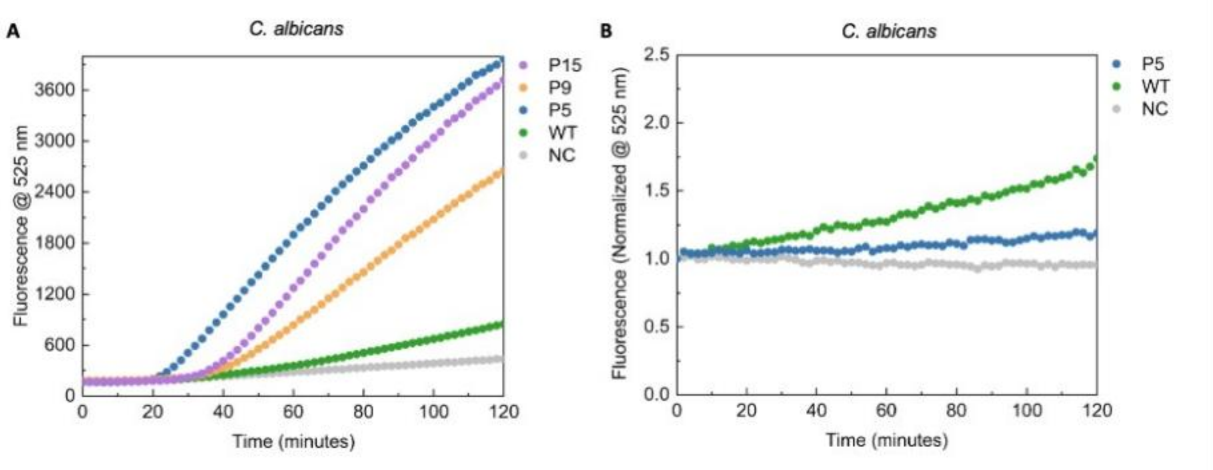


**Figure S2.** *C. albicans* engineered ITS2_3 gRNAs compared to wildtype ITS2_3 gRNA. (A) Fluorescence intensity of one-pot gRNA screen over 120 minutes. (B) Fluorescence intensity of Cas12a gRNA screen over 120 minutes. The negative controls (NC) without gRNA are in grey.

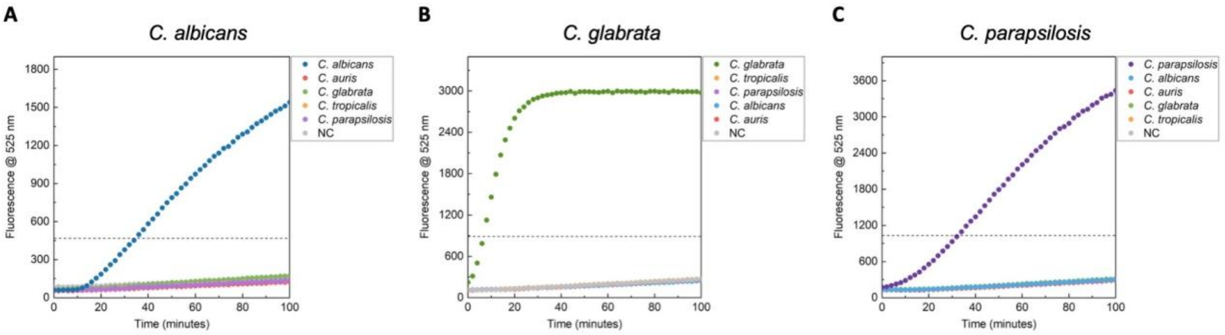


**Figure S3. *Candida* clinical isolate CRISPR-Cas12a one-pot assays.** (A) Fluorescence intensity of *C. albicans* detection, (B) *C. glabrata* detection (green) and (C) *C. parapsilosis* detection (purple) over 100 minutes. DNA was extracted from *Candida* samples from Shared Hospital Laboratory (Toronto, Canada) using the easyMAG platform. The negative controls (NC) without genomic DNA are in grey. Reference lines confirming detectable signal were defined by 3 times the negative control at 120 minutes.

**Table S2. Common antifungal resistance SNPs in *Candida ERG11*, *FKS1* and *FKS2* genes**. SNP nomenclature refers to the wildtype amino acid, followed by its position, and then the substituted amino acid (i.e. Y132F). Highlighted in red are the AMR hotspots we chose to study in this research.

| | |
|---|---|
| **Echinocandin resistance mutations in *FKS1* or *FKS2* genes** | |
| *C. albicans* | S645P, S645Y, S645F |
| *C. glabrata* | *FKS1:* S629P *FKS2*: S663P, F659Y, D666V |
| *C. parapsilosis* | P660A, S656P |
| **Azole resistance mutations in *ERG11* gene** | |
| *C. albicans* | Y132F, K143R, F126L |
| *C. glabrata* | C108G, C423T, Y141H, S410F |
| *C. parapsilosis* | Y132F |

**Table S3. gRNA and RPA primer designs for identifying *Candida* AMR mutations.** PAM sites highlighted in bold correspond to the first four nucleotides of each sequence. The capitalized nucleotides represent the amino acid codons of interest. The nucleotides in red represent the location of the SNP mutation. Nucleotides in blue represent the engineered synthetic mismatch. All sequences are listed in the 5' to 3' orientation. gRNAs were designed in the RNA form of these sequences from 5' to 3', made up of the 20 nucleotides downstream of the PAM site. Primer sequences were designed as DNA with lengths between 30 – 35 bp long, and amplicons ≥ 100 bp.

| ***C. albicans*** | ***ERG11* target sequence** | **gRNA sequence** |
|---|---|---|
| gRNA 126 | ttaactactccagttttcgg | **ttta**ccGA**A**aacuggaguaguuaa |
| gRNA 126-E | ttaactactccagttttcgg | **ttta**ccGA**A**a**u**cuggaguaguuaa |
| gRNA 132 | aggggttatttatgattgtc | **tttg**gacaaucA**U**Aaauaaccccu |
| gRNA 132-E | aggggttatttatgattgtc | **tttg**gacaau**g**A**U**Aaauaaccccu |
| ***C. glabrata*** | ***FKS2* target sequence** | **gRNA sequence** |
| gRNA FKS2 | ctgatagggtctcttagaga | **tttg****U**CUcuaagaG**A**Cccuaucag |
| gRNA E1 | ctgatagggtctcttagaga | **tttg****U**C**A**cuaag**u**G**A**Cccuaucag |
| gRNA E2 | ctgatagggtctcttagaga | **tttg****U**C**A**cuaagaG**A**Cccuaucag |
| gRNA E3 | ctgatagggtctcttagaga | **tttg****U**CUcuaag**u**G**A**Cccuaucag |
| ***C. parapsilosis*** | ***FKS1* target sequence** | **gRNA sequence** |
| gRNA 656 | ctcaatggcaaagtcaagaa | **tttc**uucuugacuuug**C**CAuugag |
| gRNA 656-E | ctcaatggcaaagtcaagaa | **tttc**uucuugacuu**a**g**C**CAuugag |
| gRNA 660 | agatgctattagaaacttgt | **tttg**acaaguuucuaauAG**C**aucu |
| gRNA 660-E | agatgctattagaaacttgt | **tttg**acaaguuucuaau**U**G**C**aucu |
| ***C. albicans ERG11* primers** | | |
| Forward primer 2 | atctgatgtttctgctgaagatgcttataa | |
| Reverse primer 1 | ggaacatatcttttaaatgaatcagtagtc | |
| ***C. glabrata FKS2* primers** | | |
| Forward primer 2 | cttctcagactttcaccgcatcttttgccc | |
| Reverse primer 1 | ctgcattttttcttacacatgtgtttttct | |
| ***C. parapsilosis FKS1* primers** | | |

| | |
|---|---|
| Forward primer 2 | tgtcaagttaagaggattggacatgtggat |
| Reverse primer 2 | cgtacatcaatcccaacacaatcttggcttgttgt |

**Table S4.** Inferred result tables for *C. albicans*, *C. glabrata*, and *C. parapsilosis* AMR detection. 126-E and 132-E gRNAs correspond to AMR detection in the *C. albicans ERG11* gene. FKS2-E1 gRNA corresponds to AMR detection in the *C. glabrata FKS2* gene. 656-E and 660-E gRNAs correspond to AMR detection in the *C. parapsilosis FKS1* gene. The coloured columns denote possible combinations of results one could obtain for each species. The inferred result column gives the expected resistance towards the corresponding antifungal. The last column shows the level of analytical sensitivity displayed by the gRNAs.

| | | | |
|---|---|---|---|
| ***C. albicans*** | | | |
| **126-E gRNA** | **132-E gRNA** | **Inferred Result** | **Sensitivity** |
| ✓ | ✓ | Neither 126 nor 132 SNPs detected. | aM |
| ✓ | ✗ | 132 SNP detected; possible azole resistance[11]. | aM |
| ✗ | ✓ | 126 SNP detected; possible azole resistance[11]. | aM |
| ✗ | ✗ | Both SNPs detected; possible azole resistance[11]. | - |
| ***C. glabrata*** | | | |
| **FKS2-E1 gRNA** | | **Inferred Result** | **Sensitivity** |
| ✓ | | Neither 663 nor 666 SNPs detected. | aM |
| ✗ | | 663 and/or 666 SNP detected; possible echinocandin resistance[3]. | - |
| ***C. parapsilosis*** | | | |
| **656-E gRNA** | **660-E gRNA** | **Inferred Result** | **Sensitivity** |
| ✓ | ✓ | Both SNPs detected; reduced echinocandin susceptibility[61] and pan-echinocandin resistance[62]. | aM |
| ✓ | ✗ | S656P SNP detected; pan-echinocandin resistance[62]. | aM |
| ✗ | ✓ | P660A SNP detected; reduced echinocandin susceptibility[61]. | aM |
| ✗ | ✗ | Neither S656P nor P660A SNPs detected. | - |

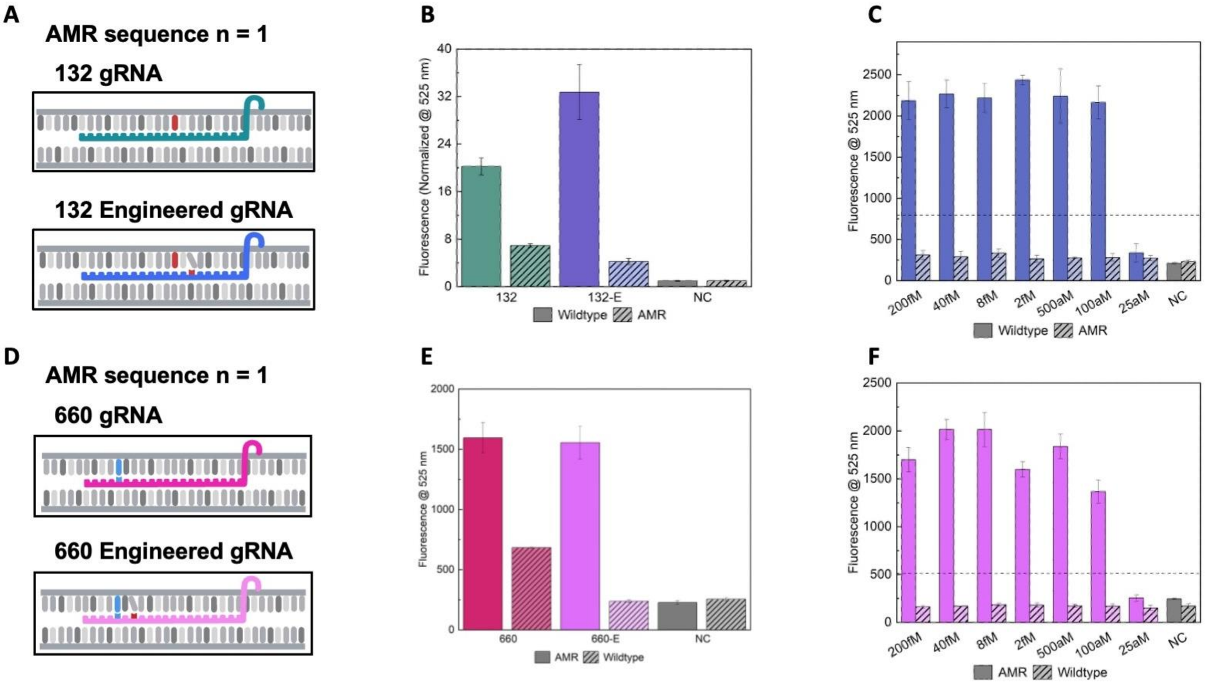


**Figure S4. *C. albicans* 132 AMR hotspot detection in the *ERG11* gene and *C. parapsilosis* 660 AMR hotspot detection in the *FKS1* gene.** (A) 132 wildtype gRNA (green) and 132 engineered gRNA (blue) designs with introduced synthetic mismatches. The red nucleotide in the gRNA represents a synthetic mismatch with the shifted nucleotide, and the red nucleotide in the AMR sequence represents a SNP. (B) Fluorescence intensity of two-pot 132 gRNA screen after 120 minutes. Fluorescence was normalized to t = 0. The negative controls (NC) without gRNA are in grey. (C) Fluorescence intensity of one-pot sensitivity screen with gRNA 132-E, after 120 minutes. The negative controls (NC) without synthetic gBlock are in grey. Reference lines confirming detectable signal were defined by 3 times the negative control at 120 minutes. (D) 660 wildtype gRNA (magenta) and 660 engineered gRNA (pink) designs. The red nucleotide in the gRNA represents a synthetic mismatch with the shifted nucleotide. The blue nucleotide in the gRNA represents complementarity with the SNP, represented by the blue nucleotide in the AMR sequence. (E) Fluorescence intensity of one-pot 660 gRNA screen after 120 minutes. The negative controls (NC) without gRNA are in grey. (F) Fluorescence intensity of one-pot 660 AMR hotspot sensitivity screen with gRNA 660-E, after 120 minutes. The negative controls (NC) without synthetic gBlock are in grey. Reference lines confirming detectable signal were defined by 3 times the negative control at 120 minutes.

**Table S5. Target sequences for CRISPR-Cas12a species detection**. Sequences were ordered on IDT as 500 bp gBlock fragments and rehydrated to 10 nM. Sequences are ordered from 5' to 3'.

| ***Candida* species** | **Internal transcribed spacer (*ITS*) target sequence** |
|---|---|
| *C. albicans* | tccgtaggtgaacctgcggaaggatcattactgatttgcttaattgcaccacatgtgtttttctttgaaacaaacttgctttggcggtgggcccagcctgccgccagaggtctaaacttacaaccaatttttttatcaacttgtcacaccagattattactaatagtcaaaactttcaacaacggatctcttaatatgaattgcagatattcgtgaatcatcgaatctttgaacgcacattgcgccctctggtattccggagggcatgcctgtttgagcgtcgtttctccctcaaaccgctgggtttggtgttgagcaatacgacttgggtttgcttgaaagacggtagtggtaaggcgggatcgctttgacaatggcttaggtctaaccaaaaacattgcttgcggcggtaacgtccaccacgtatatcttcaaactttgacctgctgaacttaagcatatcaataagcggagga |
| *C. glabrata* | tattccaaaggaggtgttttatcacacgactcgacactttctaattactacacacagtggagtttactttactactattcttttgttcgttagttctcccagtggatgcaaacacaaacaaatatttttttaaactaattcagtcaacacaagatttcttttagtagaaaacaacttcaaaactttcaacaatggatctcttggttctcgcatcgatgaagaacgcagcgaaatgcgatacgtaatgtgaattgcagaattccgtgaatcatcgaatctttgaacgcacattgcgccctctggtattcatgcctgtttgagcgtcatttccttctcaaacacgttgtgtttggtagtgagtgatactctcgtttttgagttaacttgaaattgtaggccatatcagtatgtgggacacgagcgcaagcttctctattaatctgctgctcgtttgcgcgatatta |
| *C. parapsilosis* | tatggaagtaaaagtcgtaacaaggtttccgtaggtgaacctgcggaaggatcattacagaatgaaaagtgcttaactgcatttttttcttacacatgtgtttttctttttttgaaaactttgctttggtaggccttctatatggggcctgccagagattgaactcaaccaaattttatttaatgtcaaccgattatttaatagtcaaaactttcaacaacggatctcttggttctcgcatcgatgaagaacgcagcgaaatgcgataagtaatatgaattgcagatattcgtgaatcatcgaatctttgaacgcacattatgcctgtttgagcgtcatttctccctcaaaccctcgggtttggtgttgagcgatacgctgggtttgcttgaaagaaaggcggagtataaactaatggataggtttttccactcattggtacaaactccaaaacttcttccaaattcgacctcaa |

**Table S6. Target sequences for CRISPR-Cas12a AMR detection.** Sequences were ordered on IDT as 500 bp gBlock fragments and rehydrated to 10 nM. Sequences are ordered from 5' to 3'.

| **AMR hotspot** | **Target sequence** |
|---|---|
| *C. albicans ERG11* (wildtype) | tggccgatagagctccgtgggattccttggggttctgcagcttcatatggtcaacaacccgaatcatgtcgtcggtgatggggggacggggtccggtcatggtttttaatgctaaattatctgatgtttctgctgaagatgcttataaacatttaactactccagttttcggtaaaggggttatttatgattgtccaaattccagattaatggaacaaaaaaaatttgctaaatttgctttgactactgattcatttaaaagatatgttcctaagattagagaaggctgctcatggggttgccaatggctcaaccagcactgcttcaagggtgatggagaaggaccgttca |
| *C. albicans ERG11* (AMR mutant) | gcgccggcgcggccgcgctggccgatagagctccgtgggattccttggggttctgcagcttcatatggtcaacaacccgaatcatgtcgtcggtgatggggggacggggtccggtcatggtttttaatgctaaattatctgatgtttctgctgaagatgcttataaacatttaactactccagttctcggtaaaggggttatttttgattgtccaaattccagattaatggaacaaaaaaaatttgctaaatttgctttgactactgattcatttaaaagatatgttcctaagattagagaaggctgctcatggggttgccaatggctcaaccagcactgcttcaagggtgatggagaaggaccgttca |
| *C. glabrata FKS2* (wildtype) | gccactgttttattcttctcgattatgccattaggtggtcttttcacctcatatatgcaaaaatcaagtagaagatatgttgcttctcagactttcaccgcatcttttgccccattacaaggtttggatagatggttatcttatttagtttgggttacagttttgctgccaaatactctgaatcgtacttcttcttgattttgtctctaagagaccctatcagaattttatcaactactaccatgagatgtactggtgagtattggtggggttcaaagttatgtagacatcaatcgaagattgttttaggtttcatgattgctacagatttcattttgttcttccttgatacttatttgtggtacattgttgtcaacactgtcttctctgttggtaaatccttctatctaggtatttccatct |
| *C. glabrata FKS2* (663 & 666 SNP) | gccactgttttattcttctcgattatgccattaggtggtcttttcacctcatatatgcaaaaatcaagtagaagatatgttgcttctcagactttcaccgcatcttttgccccattacaaggtttggatagatggttatcttatttagtttgggttacagttttgctgccaaatactctgaatcgtacttcttcttgattttgcctctaagagtccctatcagaattttatcaactactaccatgagatgtactggtgagtattggtggggttcaaagttatgtagacatcaatcgaagattgttttaggtttcatgattgctacagatttcattttgttcttccttgatacttatttgtggtacattgttgtcaacactgtcttctctgttggtaaatccttctatctaggtatttccatct |
| *C. glabrata FKS2* (663 SNP) | gccactgttttattcttctcgattatgccattaggtggtcttttcacctcatatatgcaaaaatcaagtagaagatatgttgcttctcagactttcaccgcatcttttgccccattacaaggtttggatagatggttatcttatttagtttgggttacagttttgctgccaaatactctgaatcgtacttcttcttgattttgcctctaagagaccctatcagaattttatcaactactaccatgagatgtactggtgagtattggtggggttcaaagttatgtagacatcaatcgaagattgttttaggtttcatgattgctacagatttcattttgttcttccttgatacttatttgtggtacattgttgtcaacactgtcttctctgttggtaaatccttctatctaggtatttccatct |
| *C. glabrata FKS2* (666 SNP) | gccactgttttattcttctcgattatgccattaggtggtcttttcacctcatatatgcaaaaatcaagtagaagatatgttgcttctcagactttcaccgcatcttttgccccattacaaggtttggatagatggttatcttatttagtttgggttacagttttgctgccaaatactctgaatcgtacttcttcttgattttgtctctaagagtccctatcagaattttatcaactactaccatgagatgtactggtgagtattggtggggttcaaagttatgtagacatcaatcgaagattgttttaggtttcatgattgctacagatttcattttgttcttccttgatacttatttgtggtacattgttgtcaacactgtcttctctgttggtaaatccttctatctaggtatttccatct |
| *C. parapsilosis* | tctgttgttggattcttcattgctgttgcaactttggttttctttgctgtcatgcctttgggaggtttgttcacctcgtatatgaacaagaggtcgaggaggtacatctcgtctcatacttttactgcaaactttgtcaagttaagaggattggacatgtggatgtcgtatttgttgtgggtcttggttttcctagcaaagttggttgaatcttatttcttcttgactttgtcattgagagatcctattagaaacttgtcaaagaccacaatgagatgtaccggtgaagtttggtatggtgacattgtctgtagacaacaa |

| | |
|---|---|
| *FKS1* (wildtype) | gccaagattgtgttgggattgatgtacgctgtcgatttgttgttgttctttttggatacctacttgtggtacattatctgtaattgtatcttttccattggtcgttcattctatttgggtatctcaatcttgacaccatggagaaacatcttt |
| *C. parapsilosis FKS1* (AMR mutant) | tctgttgttggattcttcattgctgttgcaactttggttttctttgctgtcatgcctttgggaggtttgttcacctcgtatatgaacaagaggtcgaggaggtacatctcgtctcatacttttactgcaaactttgtcaagttaagaggattggacatgtggatgtcgtatttgttgtgggtcttggttttcctagcaaagttggttgaatcttatttcttcttgactttgccattgagagatgctattagaaacttgtcaaagaccacaatgagatgtaccggtgaagtttggtatggtgacattgtctgtagacaacaagccaagattgtgttgggattgatgtacgctgtcgatttgttgttgttctttttggatacctacttgtggtacattatctgtaattgtatcttttccattggtcgttcattctatttgggtatctcaatcttgacaccatggagaaacatcttt |

**Table S7. *Candida* clinical isolates.** Extracted DNA fungal samples were collected by Dr. Rob Kozak from Shared Hospital Laboratory (Toronto, Canada).

| **Sample** | ***Candida* species** | **Concentration (ng/µL)** |
|---|---|---|
| FUN_001 | *C. tropicalis* | 1.085 |
| FUN_002 | *C. parapsilosis* | 1.690 |
| FUN_005 | *C. glabrata* | 0.895 |
| FUN_006 | *C. albicans* | 6.445 |
| FUN_008 | *C. albicans* | 1.310 |
| FUN_009 | *C. albicans* | 2.720 |
| FUN_010 | *C. glabrata* | 4.585 |
| FUN_012 | *C. albicans* | 1.995 |
| FUN_013 | *C. parapsilosis* | 0.320 |
| FUN_014 | *C. auris* | 1.465 |
